# Salivary CGRP as diagnostic and migraine attack phase monitor biomarker: CGRP (in)dependent attacks

**DOI:** 10.1101/2020.11.18.20233841

**Authors:** Alicia Alpuente, Victor J Gallardo, Laila Asskour, Edoardo Caronna, Marta Torres-Ferrus, Patricia Pozo-Rosich

## Abstract

**Objective:** To assess saliva as a substrate to measure CGRP by comparing interictal levels of CGRP in patients with episodic migraine and controls; and to evaluate saliva CGRP temporal profile during migraine attacks.

**Methods:** This prospective observational study included women with episodic migraine and healthy controls. Participants collected daily saliva samples for 30 consecutive days and 3 additional ones during migraine attacks. 4 timepoints were considered: interictal (72h headache-free), preictal (PRE-24h before the attack), ictal (0h,2h,8h), postictal (POST-24h after the attack). CGRP levels were quantified by ELISA.

**Results:** 35 women (22 patients, 13 controls) were included. Statistically significant differences were found in interictal salivary levels of CGRP between patients and controls (median[IQR]: 98.0 [86.7] vs. 42.2 [44.7] pg/mL; p=0.010). An increase in CGRP levels during migraine attacks was detected (median[IQR]: preictal 113.5 [137.8], 0h 164.6 [204.5], 2h 101.7 [159.1], 8h 82.6 [166.2], postictal 79.6 [124.3] pg/ml; p<0.001). Patients were classified as having CGRP-dependent (80.0%) and non-CGRP dependent migraine attacks (20.0%) according to the magnitude of change between preictal and ictal phase (0h). Accompanying symptoms were different depending on the type of attack. In the longitudinal analysis, we observed that the amount of CGRP measured during attacks were phase dependent and it was influenced by the frequency of monthly headache days (p=0.02).

**Interpretation:** Patients with episodic migraine have higher interictal salivary levels of CGRP than controls. These levels usually increase during a migraine attack, however, not every attack is CGRP-dependent, which in turn, might explain different underline pathophysiology and response to acute and preventive treatment.

## INTRODUCTION

Biomarkers are indicators of a normal biological or pathological process and they have great value in clinical practice and research ^1^. Being migraine the most prevalent and disabling neurological disease ^2^, specific biomarkers have yet not been validated ^3,4^, reflecting the dynamism and complexity of the disease. However, the most promising candidate of becoming a migraine biomarker is currently calcitonin gene related peptide (CGRP) due to its clear implication in migraine pathophysiology ^5^, underpinned by the arrival of therapies targeting CGRP pathways ^6^.

Saliva as substrate to study biomarkers is a worthwhile approach because its collection is noninvasive, inexpensive and neuropeptides could be monitored because samples can be repeatedly obtained from subjects ^7^. The feasibility of CGRP detection in human saliva has been previously demonstrated and used as a way to study trigeminovascular activation in migraine ^8-13^. CGRP has been considered as a potential diagnostic ^8,14,15^ and treatment predictive biomarker in acute and preventive migraine treatment ^13,15-17^. Nonetheless, we found a large variability between CGRP levels across studies, probably due to methodological differences such as saliva collection techniques, type of assay and heterogeneous study population, making the results not comparable.

Biomarker assay validation before introduction in clinical practice is essential in order to get reliable results and assay information is mandatory in research reports on novel biomarker candidates ^18^. A standardized approach to saliva collection and analysis is needed in order to be able to use it as a valuable resource to gain scientific and diagnostic information in migraine. Therefore, the objectives of this study were: 1) to define a highly specific technical protocol to measure salivary CGRP and, 2) to compare interictal salivary CGRP levels in episodic migraine (EM) patients with healthy controls (HC), as well as, 3) to assess the temporal profile of CGRP during migraine attacks.

## METHODS

### Participants and study design

This is a prospective longitudinal exploratory study. Participants were recruited from the outpatient headache clinic and carefully interviewed by a headache specialist. Recruitment period was from March 2018 to November 2019.

In order to obtain a homogeneous sample, HC were women between 18-65 years old with no previous migraine or headache history. Patients fulfilled the criteria for EM with or without aura, according to the International Classification of Headache Disorders (ICHD-3) ^19^. Participants with a smoking habit; medical diagnosis of anxiety/depression; medical diagnosis of presence chronic pain disorders; subjects taking medication affecting central nervous system; subjects taking migraine preventive medication in the past year prior to the study or history of any medical condition that could alter saliva content were excluded.

At the screening visit, demographical and clinical data were collected. All participants completed the following Patient Reported Outcome Scales (PROs): Migraine Disability Assessment (MIDAS) ^20^ and the Headache Impact Test 6 (HIT-6) ^21^. Patients also completed the Hospital Anxiety and Depression Scale (HAD A-D) ^22^, Perceived Stress Scale (PSS) ^23^,12-item Allodynia Symptom Checklist (ASC-12) ^24^, The International Physical Activity Questionnaire (IPAQ), The Pittsburgh Sleep Quality Index (PSQI) ^25^ and a PERIODONTAL assessment.

All participants were given detailed verbal and written instructions for saliva collection and they were provided with appropriate material for its collection at home including: pre-labelled tubes; diaries to register sample collection time and menstrual cycle; questionnaires to record migraine attack characteristics such as pain intensity and duration, accompanying symptoms and acute treatment used. During the study, subjects treated their migraine attacks as usual, if they considered they could not tolerate it, with an approval of the investigator at the initial visit (triptans and non-steroidal anti-inflammatory drugs were allowed).

### Saliva collection

HC and EM patients collected saliva samples consecutively during a 30-day period (baseline samples). EM patients collected three extra saliva samples in each migraine attack (painful condition samples): 0h, 2h and 8h.

Saliva collection was carried out with the resting unstimulated whole saliva method ^7,11^ using the following step-by-step indications:

- To collect saliva at the same time of the day, early in the morning, fasting condition.
- Not to eat, drink or brush their teeth before collection.
- To rinse their mouth with water, discarding initial saliva in order to avoid contaminated saliva with debris.
- To collect the fluid by spitting into a sterile tube of 5 mL for 5 minutes with a minimum quantity of 3 mL. Each tube was used once only.
- Not to use citric acid since it could degrade CGRP.

After collection, all saliva samples were kept in the freezer of participants at −20°C until they were brought in to the laboratory. During transport samples were kept on ice. We kindly asked participants to keep the attack samples in a fridge if they were away from home. Once saliva samples were received at the laboratory they were stored in the freezer at −20ºC.

### Plasma collection

Plasma samples were also collected from each participant on day 1 in order to make a saliva-plasma correlation. It was noted if it was a free pain period (outside of an attack). Samples were collected from the subject’s antecubital vein between 8-10 am and transferred to EDTA coated 10.8 mg tubes (BD Vacutainer System, K2E). All samples were centrifuged at 3500 rpm at 4ºC for 15 min and supernatants were immediately stored at −80ºC until analysis.

### Saliva extraction

Saliva samples were thawed slowly at room temperature and then placed on ice. The samples were centrifuged for 20 minutes at 3500 rpm-4ºC, and the supernatant was aliquoted into 1.5mL centrifuge sterile tubes and stored at −80ºC or immediately analyzed. Prior to Enzyme-linked Immunosorbent Assay (ELISA), samples were thawed slowly and then centrifuged for 5 minutes at 3500 rpm at room temperature to pellet cellular debris.

Samples were stored at −80ºC, which helped avoid protein degradation and denaturalization. Moreover, to minimize degradation of unstable antigens, samples were kept in ice, and, repeated freeze-thaw cycles were avoided.

### Assay protocol

The operating procedures for assay performance are exhaustively detailed in assay protocol. CGRP was quantitated using a commercial CGRP sandwich ELISA kit (CUSABIO®, CSB-E08210H, detection range: 1.56–100 pg/ml, minimal detectable dose: 0.39 pg/ml), following manufacturer’s instructions (Wuhan, China). Intra-assay precision and inter-assay precision is declared with a coefficient of variation (CV) of<8%, respectively<10%. Duplicate measurements were performed for each sample. CGRP concentrations were determined from calibration curves using a 4PL fitting as implemented in Analysis software Gen 5 resulting in a fit with R2>0.99 in every case. The final CGRP level of each sample was calculated as the average of the two measurements. An internal validation of the test was performed, ensuring that the ELISA assay used was reliable (quality control was included in the kit).

We found that saliva samples had to be diluted 1:10 with sample diluent, being important to homogenize the sample with the diluent (repeated up-down cycles). In addition, it is extremely important to discard blood contaminated samples. In plasma samples, it is important to discard lipemic and hemolyzed samples. Those samples were not diluted due to the low concentration of CGRP in plasma.

CGRP measurements were performed at our laboratory at the Vall d’Hebron Institute of Research, Spain. The immunoassay used did not require CGRP values to be normalized according to saliva volume and amount of total protein but CGRP concentration was corrected by inter and intra-assay coefficients of variability.

In order to analyze CGRP levels over the different migraine phases, we considered 4 timepoints: interictal (median value of 5 consecutive days, for migraine patients when they were headache-free for 72 hours), preictal (pre-24h before the migraine attack), ictal (0h,2h,8h) and postictal (post-24h after the migraine attack).

### Statistics

Descriptive and frequency statistics were obtained and comparisons made using R v3.6.3. Nominal (categorical) variables were reported as frequencies (percentages) while mean±standard deviation (age, disease evolution time, HIT-6 and PSS) or median and interquartile range (Me[IQR]) (MIDAS, HAD Scale and CGRP levels) were reported for continuous variables. Normality assumption of quantitative variables was checked through visual methods (Q-Q plots) and normality tests (Shapiro-Wilk test).

Statistical significance for intergroup variables was assessed by Pearson’s chi-square when comparing categorical variables. In the case of having an expected count less than 5 in more than 20% of cells in the contingency table, Fisher’s exact test was used. Linear trend chi-square was considered for ordinal variables. Independent t-test for continuous variables that followed a normal distribution (age and PSS) was used in order to assess differences between migraine patients and healthy controls and, Mann-Whitney U test was used for the rest variables that did not follow any normality assumption (HAD Scales and CGRP basal levels). The degree of association between interictal CGRP levels and clinical variables was computed by Spearman’s rank correlation and summarized by Spearman’s rho coefficient and related p-values.

We estimated the ictal log2Fold-Change (*log*_2_*FC_ictal_*) (Eq. 1) in patients with the aim of finding out whether all migraine attacks reflected an incremental change at salivary CGRP level between preictal phase (pre) and ictal phase (0h). Hence, migraine attacks were classified into CGRP dependent (dCGRP) (log> 0) or non-CGRP dependent (nCGRP) (*log*_2_*FC_ictal_* ≤ 0).

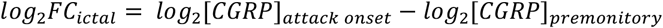

Equation 1. Ictal Fold-Change: Measure concept for the evaluation of the change of the CGRP salivary level between premonitory and the attack onset phases.

Finally, to determine the significance of CGRP intraindividual changes according to patient’s migraine phase, we used the nonparametric Friedman test. Moreover, we evaluated the interaction between the migraine phase effect in CGRP concentration with the basal headache frequency (≤5 days/month or 6-10days/month) or the use of specific acute medication category (None, non-NSAID or triptan) with nonparametric rank-based methods for longitudinal data using the nparLD R package (v2.1) ^26^. Descriptive point estimators of relative factor effect (RFE), confidence intervals and the ANOVA-type statistic (ATS) are reported. Nonparametric tests were chosen due to small sample sizes and because CGRP levels did not followed a normal distribution.

A statistical power calculation was not conducted prior to the study because the sample size was based on the available data. P-values presented are for a two-tailed test and p-values <0.05 were considered statistically significant.

Approval for this study was obtained from the Vall d’Hebron Ethics Committee (PR(IR)292/2017). All participants gave their consent for data collection.

## RESULTS

### Descriptive

In total, 22 EM and 13 HC with a mean age of 27.9±8.9 years old were included (table 1). EM patients had statistically significant higher proportion of perceived stress (95.5% vs. 46.2%; p=0.002). Twelve patients (54.5%) reported headache frequency of ≤5 days/month and 45.4% (10/22) reported headache frequency of 6-10 days/month.

**Table 1.** Baseline participants demographics, clinical characteristics and comorbidities.

|  |  | HC<br>(n=13) | EM<br>(n=22) | P value |
| --- | --- | --- | --- | --- |
| Demographics and lifestyle |  |  |  |  |
| Age, mean±SD, years old |  | 28.9±6.0 | 30.4±9.4 | 0.156a |
| Physical Activity, n (%) | Low | 5 (41.7%) | 6 (31.6%) | 0.365b |
|  | Medium | 5 (41.7%) | 12 (63.2%) |  |
|  | High | 2 (16.7%) | 1 (5.3%) |  |
| Work shift, n (%) | Early | 7 (58.3%) | 13 (68.4%) | 0.705c |
|  | Late | 5 (41.7%) | 6 (31.6%) |  |
| Migraine characteristics |  |  |  |  |
| Basal Headache Frequency, n (%) | ≤5d/mo |  | 12 (54.5%) |  |
|  | 6-10d/mo |  | 10 (45.5%) |  |
| Evolution time, mean±SD, years |  |  | 13.4±11.0 |  |
| Aura, n (%) |  |  | 8 (36.4%) |  |
| MIDAS, median [IQR] |  |  | 20.0 [49.8] |  |
| HIT-6, mean±SD |  |  | 63.7±5.9 |  |
| Comorbidities |  |  |  |  |
| Anxiety (HADS ≥ 8), n (%) |  | 5 (38.5%) | 12 (54.5%) | 0.489c |
| Depression, (HADS ≥ 8), n (%) |  | 1 (7.7%) | 4 (18.2%) | 0.274c |
| Perceived stress (PSS ≥ 14), n (%) |  | 6 (46.2%) | 21 (95.5%) | 0.002* |

HC: Healthy Control; EM: Episodic Migraine; SD: Standard Deviation; IQR: Interquartile Range; HADS: Hospital Anxiety and Depression Scale; PSS: Perceived Stress Scale; d/mo: days/month.
aIndependent T-test
bLinear trend chi-squared
cFisher's exact test
\*P value ≤ 0.01

### Interictal CGRP levels

We found statistically significant higher interictal CGRP-like immunoreactivity (CGRP-LI) salivary levels when comparing EM and mean basal HC values (98.0[86.7] vs. 42.2[44.7] pg/mL; p=0.010) (Fig. 1A). We did not find any correlation between CGRP-LI levels in regards to: age (rho=0.281, p=0.102), HADS-A (rho=0.282, p=0.101), HADS-D (rho=0.319, p=0.062), or PSS score (rho=0.243, p=0.159). Although plasma CGRP-LI levels in EM patients were higher than HC, we did not find statistically significant differences (EM: 6.1[5.0] vs. HC: 5.3[6.3] pg/mL; p=0.205) (Fig. 1B). There was not a correlation between salivary and plasma CGRP-LI levels.

**Figure 1.**
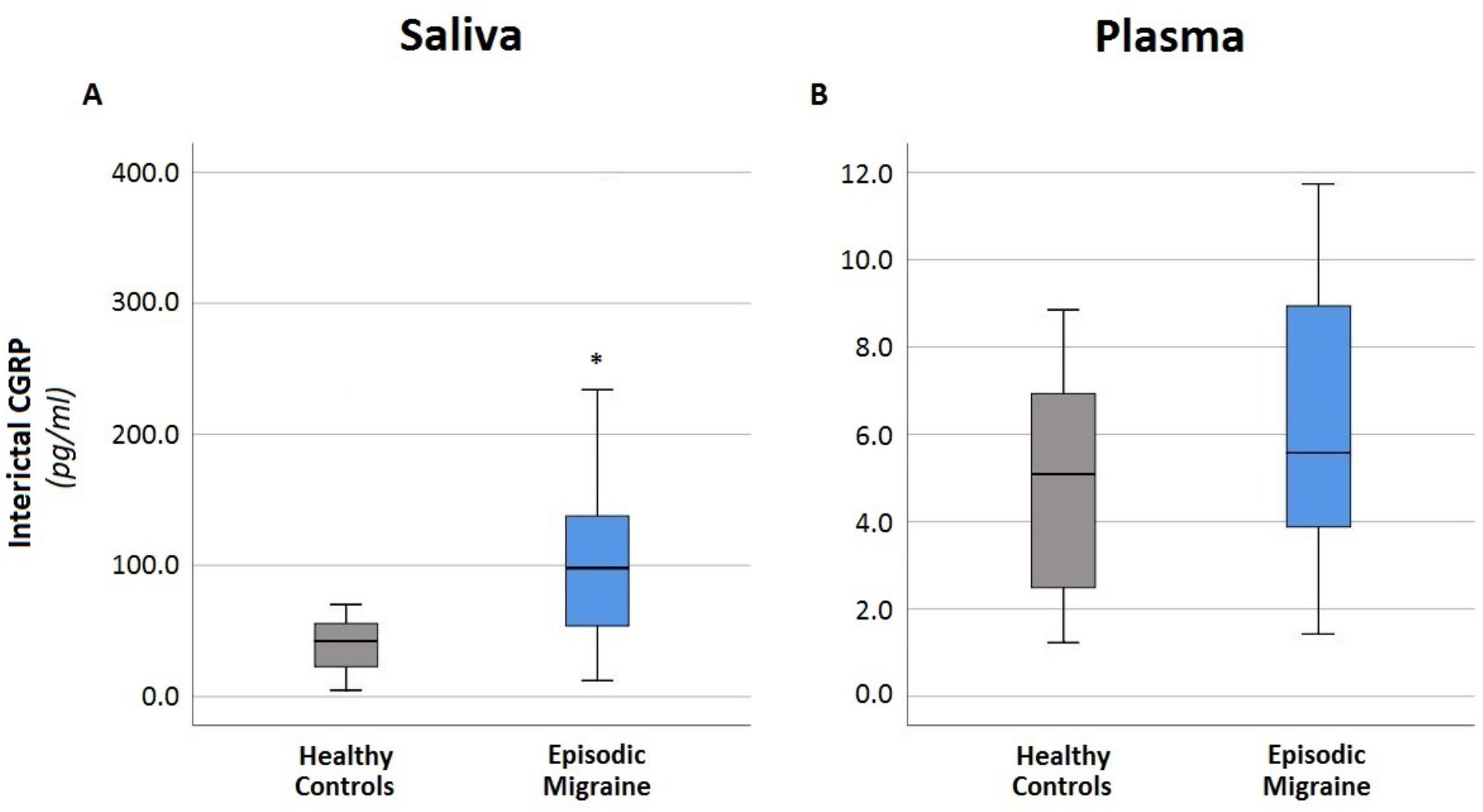
Interictal salivary levels of CGRP. Box plots and 95% confidence intervals for interictal CGRP levels in saliva (A) and plasma (B). Interictal salivary levels of CGRP were calculated as the median value of 5 consecutive random days in healthy controls. In migraine patients, interictal salivary CGRP levels were calculated as the median value of 5 consecutive days when they were headache-free for 72 hours (interictal period). Plasma CGRP values were extracted during the screening, patients did not report a migraine attack during plasma blood extraction. *p value ≤ 0.01

### Longitudinal analysis: salivary CGRP levels through migraine attack

Amongst all patients, a total of 40 migraine attacks were collected. CGRP-LI concentration in saliva changed in a statistically significant way, in each timepoint considered of the attack, when analyzing all migraine attacks (pre:113.5[137.8], ictal:164.6[204.5], 2h:101.7[159.1], 8h:82.6[166.2], post:79.6[124.3] pg/ml; p<0.001) (Fig. 2).

**Figure 2.**
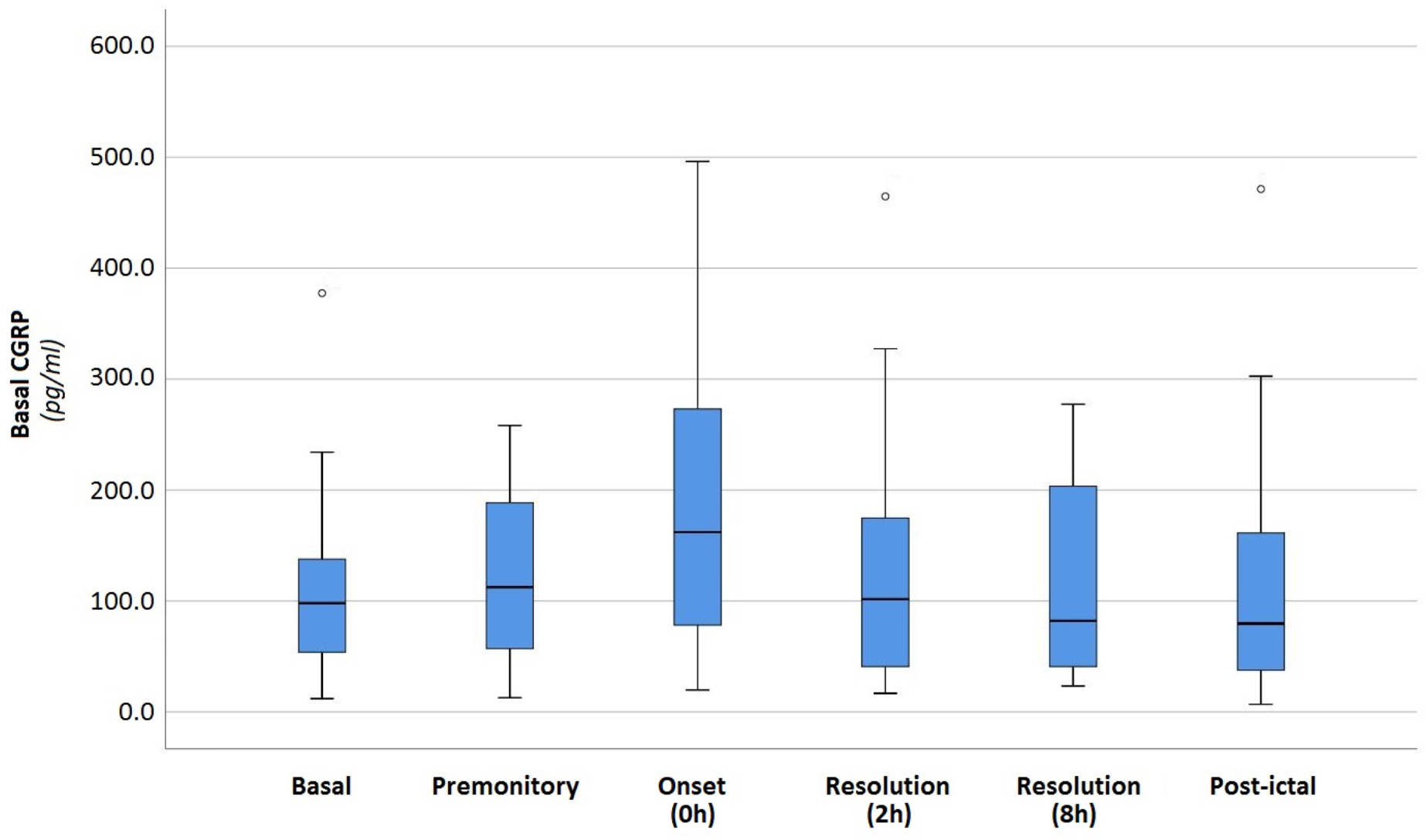
CGRP values through the different migraine phases. Box plots and 95% confidence intervals for CGRP levels evolution in saliva according to patients’ migraine phase.

When we analyzed the interaction between the migraine phase effect and headache frequency, we found that participants who reported headache frequency ≤5 days/month had statistically significant smaller magnitude of change in CGRP-LI levels over the different migraine phases compared to those who reported 6-10 days/month (ATS p=0.020) (Fig. 3A). Nonetheless, this interaction did not reach the level of statistical significance between medicated attacks with neither non-steroidal anti-inflammatory drugs (NSAID) nor triptans even though it showed smaller magnitude of change in CGRP-LI levels in triptan-treated attacks after migraine attack onset (ATS p=0.436) (Fig. 3B).

**Figure 3.**
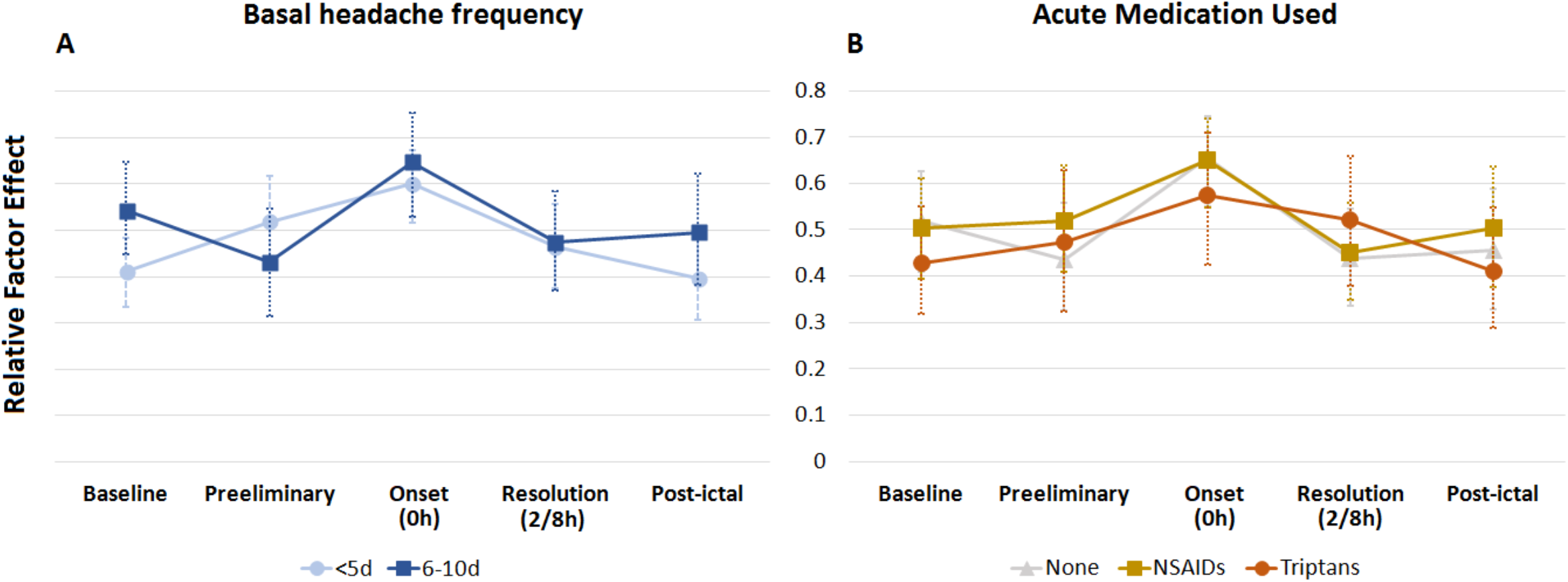
Interaction between the evolution in CGRP levels in the different migraine phases and (A) basal headache frequency or (B) acute treatment for migraine attacks. We studied the interaction between the migraine phase effect and the basal headache frequency (A) and acute treatment (B) using rank-based nonparametric methods for longitudinal data. Only the interaction in the evolution of CGRP levels between the migraine phase and basal headache frequency was statistically significant.

### Subtypes of migraine patients

We found that 80.0% (32/40) were CGRP dependent (dCGRP) and 20.0% (8/40) were non-CGRP dependent (nCGRP). When we compared interictal CGRP-LI levels between these two groups, no statistically significant differences were found; except for the onset of migraine attack (0h), CGRP dependent group presented a statistically significant higher levels of CGRP-LI (dCGRP: 172.4 [252.1] vs. nCGRP: 60.9 [77.0]; p<0.0001). In regards to accompanying symptoms, a statistically significant association between photophobia (dCGRP: 81.3% vs. nCGRP: 37.5%; p=0.025) and phonophobia (dCGRP: 75.0% vs. nCGRP: 25.0%; p=0.014) was found in dCGRP group. Dizziness (dCGRP: 34.4% vs. nCGRP: 75.0%; p=0.047) and vomiting (dCGRP: 3.1% vs. nCGRP: 37.5%; p=0.020) were statistically significantly associated with nCGRP group (table 2). When we analyzed migraine patients, 15 out of 22 patients only showed dCGRP migraine attacks; 4 out of 22 patients only showed nCGRP migraine attacks and only 3 patients showed both type of migraine attacks.

**Table 2.** Clinical parameters associated with dichotomic migraine attacks: CGRP dependent and non-CGRP dependent.

|  | Non-CGRP<br>dependent<br>(n=8) | CGRP<br>dependent<br>(n=32) | P value |
| --- | --- | --- | --- |
| CGRP (Interictal)<sup>a</sup>, median [IQR] | 60.4 [87.3] | 110.7 [83.6] | 0.127 |
| CGRP (Onset)<sup>a</sup>, median [IQR] | 60.9 [77.0] | 172.4 [252.1] | <0.0001* |
| Treated attack<sup>b</sup>, n (%) | 6 (75.0%) | 17 (53.1%) | 0.214 |
| Aura<sup>b</sup>, n (%) | 3 (37.5%) | 5 (15.6%) | 0.320 |
| Unilateral Pain<sup>b</sup>, n (%) | 1 (12.5%) | 8 (25.0%) | 0.655 |
| Nausea<sup>b</sup>, n (%) | 5 (62.5%) | 22 (68.8%) | 1.000 |
| Dizziness<sup>b</sup>, n (%) | 6 (75.0%) | 11 (34.4%) | 0.047* |
| Vomiting<sup>b</sup>, n (%) | 3 (37.5%) | 1 (3.1%) | 0.020* |
| Photophobia<sup>b</sup>, n (%) | 3 (37.5%) | 26 (81.3%) | 0.025* |
| Phonophobia<sup>b</sup>, n (%) | 2 (25.0%) | 24 (75.9%) | 0.014* |
| Allodynia<sup>b</sup>, n (%) | 6 (75.0%) | 19 (59.4%) | 0.686 |

<sup>a</sup>Mann-Whitney U test
<sup>b</sup>Fisher's exact test
\*P value ≤ 0.05

## DISCUSSION

In this study, we performed an ELISA assay to measure and monitor salivary levels of CGRP in patients with very episodic and naïve migraine patients over the different migraine phases. We wanted to clearly see how attacks started and stopped, and avoid continuums of head pain. Compared to HC, EM patients showed higher interictal CGRP levels. However, intraindividually CGRP levels change during a migraine attack. Moreover, EM patients with clear migraine clinical symptoms can or can’t have CGRP dependent attacks, creating subtypes of migraine patients, those who release CGRP during an attack and those who do not.

First, our results show that interictal salivary levels of CGRP are significantly higher in patients with EM, even if they suffer from infrequent attacks, compared to HC. Bellamy et al. also found that patients with EM had elevated salivary levels of CGRP between attacks compared to controls.^13^ There is only one previous study that found higher CGRP levels in HC probably due to participant’s selection and procedures ^8^. Other than saliva, previous studies using other substrates such as plasma ^27^ or tear fluid ^28^ also showed higher interictal levels of CGRP in patients with EM. Our finding reinforces that interictal salivary levels of CGRP are elevated in migraine patients, supporting its use as possible migraine diagnostic biomarker.

As a cycling brain disorder, monitoring migraine over its different phases is important in order to understand the underlying mechanisms and pathogenesis ^29^, as it has been previously demonstrated in neuroimaging studies ^30^. At the molecular level, our study reveals that salivary levels of CGRP change over a migraine attack according to the timepoint analyzed: they were shown to increase as headache progressed from preictal to the ictal phase and decreased in the postictal phase at or below interictal levels. Our results are the first ones to see this gradual change of CGRP levels during the attack, and confirm previous studies which also showed an increase in salivary levels of CGRP during the ictal phase, interpreting it as a distant sign of trigeminovascular activation ^8,16^ This change reflects that CGRP levels may play a role as a dynamic biomarker in a disease that is not static which in turn strength the importance of monitoring migraine at different timepoints; and considering intraindividual change and not CGRP levels at a group level.

Regarding our longitudinal analysis, we observed that CGRP concentration was dependent on the migraine attack phase. We found an association between larger variations in CGRP concentration and higher headache frequency, which may support a greater activation of the trigeminovascular system in more severe forms of the disease ^31,32^. In this regard, controversial results have been published when CGRP levels were analyzed in serum samples. Cernuda-Morollón et al. ^33^ found that CM patients exhibited the highest levels of CGRP, followed by EM patients and HC, suggesting it as a diagnostic biomarker of CM. In contrast, Lee et al. ^34^ did not find any significant differences between groups. Notwithstanding and taking this together, our results show that CGRP may serve as a biomarker of disease severity.

Based on our FC analysis, we also observed different types of patients: those with CGRP dependent and non-CGRP dependent migraine attacks. Some diagnostic migraine symptoms such as photo and phonophobia were significantly related to CGRP. This could be interesting both to understand the link between CGRP and migraine symptoms ^35^, helping also to clinically predict CGRP treatment response according to the percentage of attacks with different symptoms. In this sense, it is worth mentioning the similarity found between the percentage of patients nCGRP dependent and non-responders’ patients in clinical trials ^36^. Moreover, since migraine diagnosis is currently based on clinical criteria according to the ICHD-3 ^19^, our results supports the concept of classifying migraine from a pathophysiological point of view, ruling out some difficulties encompassed when a purely clinical diagnosis is done. This information might help us to carry out precision medicine.

Measuring CGRP closer to the afferents (saliva in our study) could be more effective than in plasma and is a reliable reflection of trigeminovascular activation ^28^. We have found that levels of CGRP were higher in saliva than in plasma and there were not significant differences in plasma levels of CGRP between EM and HC. Hence, serum is not perhaps the ideal matrix to measure CGRP levels since neuropeptides are circulating in low concentrations in serum. As in previous studies, we did not find a correlation between salivary and plasma levels of CGRP ^8,13,16^. During the past decades, saliva has received growing attention as a substrate to study biomarkers in chronic pain disorders ^37^. Salivary glands are innervated by the third branch of the trigeminal nerve and therefore are closer to the trigeminovascular system. These CGRP-containing trigeminal nerves release this neuropeptide in conditions such as migraine and cluster headache ^8^.

Since the first demonstration of an increase in CGRP levels in external jugular venous blood during a migraine attack ^38^, researchers have been seeking the most adequate technique to measure CGRP levels and, controversial results have been published possibly due to methodological differences. Our study, carried out with a rigorous methodology, strengthens the use of CGRP as a molecular biomarker, helping to diagnose the disease, correlate it with disease severity and predict treatment outcomes, which in turn would help manage both physician and patient expectations. It is now clear that CGRP plays a major role in migraine pathophysiology ^39,40^, consolidated by the approval of new therapies targeting CGRP or its receptor ^41-43^.

Deep phenotyping in migraine patients is an area of growing interest. Migraine is a complex neurological disease because of patient’s heterogeneity and its underlying pathophysiology. Moreover, its dynamic character and cycling behavior with unpredictable attacks makes it more difficult to study in a static manner. For that reason, we monitored levels of CGRP through the different migraine phases and we found different types of migraine patients from a molecular point of view. Future research should be focused on finding a molecular, anatomical, genetical and physiological way of defining migraine which in turn could help to develop a pathophysiological driven classification.

The study has some limitations. First, the small size of the sample. Some patients only collected one migraine attack so we need to be cautious when classifying patients into different types. However, we really wanted to include episodic migraine patients, in order to clearly differentiate the attack from the interictal period. Finally, most of the migraine attacks were treated. Thus, the majority of our CGRP levels were measured in medicated migraine attacks, proving that differences can be found even if we treat patients.

Our study has a longitudinal approach collecting samples over a 30-day period, which in migraine is very rare. This allowed us to analyze the CGRP profile in each migraine phase and each patient in a dynamic manner. Until now, saliva’s studies published performed at patient’s home, consisted of 1-2 days of saliva collection. Moreover, our study participants were strictly selected, without any history of previous preventive treatment use, amongst other characteristics, resulting in a very homogeneous sample.

In conclusion, our data show that CGRP levels vary according to the migraine phase, differentiating two types of patients from a pathophysiological point of view. The molecular profile of migraine might allow clinicians to better phenotype patients, may predict response to treatment and helps increase our understanding of migraine pathophysiology.

## Data Availability

The datasets used and/or analyzed during the current study are available from the corresponding author on reasonable request.

## Acknowledgement

The authors thank the migraine patients who patiently followed all of our instructions for 30 days and they did not take any acute treatment until the first samples of the migraine attacks was collected.

This study was funded by a grant from Instituto de Salud Carlos III (ISCIII - PI16/01525) and Alicia Alpuente has been partially financed by a grant from the Headache Study Group of the Spanish Neurological Society 2018.

## Authors’ contributions

AA, VJG, MTF and PPR made substantial contributions to the conception and design of the study. LA made substantial contribution to the methodology and she performed the ELISA protocol. AA wrote first draft of the paper. VJG performed statistical analysis. All authors participated in acquisition, analysis and/or interpretation of data. All authors have critically revised and finally approved the version to be published.

